# Interdependencies between cellular and humoral immune responses in heterologous and homologous SARS-CoV-2 vaccination

**DOI:** 10.1101/2021.12.13.21267729

**Authors:** Moritz M. Hollstein, Lennart Münsterkötter, Michael P. Schön, Armin Bergmann, Thea M. Husar, Anna Abratis, Abass Eidizadeh, Meike Schaffrinski, Karolin Zachmann, Anne Schmitz, Jason Scott Holsapple, Hedwig Stanisz-Bogeski, Julie Schanz, Uwe Groß, Andreas Leha, Andreas E. Zautner, Moritz Schnelle, Luise Erpenbeck

## Abstract

**Background:** Homologous and heterologous SARS-CoV-2-vaccinations yield different spike protein-directed humoral and cellular immune responses. However, their interdependencies remain elusive.

**Methods:** COV-ADAPT is a prospective, observational cohort study of 417 healthcare workers who received homologous vaccination with Astra (ChAdOx1-S; AstraZeneca) or BNT (BNT162b2; Biontech/Pfizer) or heterologous vaccination with Astra/BNT. We assessed the humoral (anti-spike-RBD-IgG, neutralizing antibodies, antibody avidity) and cellular (spike-induced T cell interferon-γ release) immune response in blood samples up to 2 weeks before (T1) and 2 to 12 weeks following secondary immunization (T2).

**Findings:** Initial vaccination with Astra resulted in lower anti-spike-RBD-IgG responses compared to BNT (70±114 vs. 226±279 BAU/ml, p<0.01) at T1, whereas T cell activation did not differ significantly. Booster vaccination with BNT proved superior to Astra at T2 (anti-spike-RBD-IgG: Astra/BNT 2387±1627 and BNT/BNT 3202±2184 vs. Astra/Astra 413±461 BAU/ml, both p<0.001; spike-induced T cell interferon-*γ* release: Astra/BNT 5069±6733 and BNT/BNT 4880±7570 vs. Astra/Astra 1152±2243 mIU/ml, both p<0.001). No significant differences were detected between BNT-boostered groups at T2. For Astra, we observed no booster effect on T cell activation. We found associations between anti-spike-RBD-IgG levels (Astra/BNT and BNT/BNT) and T cell responses (Astra/Astra and Astra/BNT) from T1 to T2. There were also links between levels of anti-spike-RBD-IgG and T cell at both time points (all groups combined). All regimes yielded neutralizing antibodies and increased antibody avidity at T2.

**Interpretation:** Interdependencies between humoral and cellular immune responses differ between common SARS-CoV-2 vaccination regimes. T cell activation is unlikely to compensate for poor humoral responses.

**Funding:** Deutsche Forschungsgemeinschaft (DFG), ER723/3-1

**Research in context:** *Evidence before this study:* We searched Pubmed for papers published between 01/01/2019 and 14/05/2021 with the search terms “covid-19” combined with “vaccination” and “heterologous”, excluding “BCG”. Of the 41 papers found, none addressed the combination of ChAdOx1-S by AstraZeneca (Astra) and BNT162b2 by Biontech/Pfizer (BNT). After our study was initiated, the CombiVacS trial reported a significant booster effect when BNT was given after initial vaccination with Astra.^1^ The investigators of the CoCo trial subsequently published data on heterologous immunization in comparison to homologous Astra in a small population (n=87), with the heterologous immunization scheme showing a superior humoral and cellular immune response.^2^ Further studies investigated heterologous vaccinations with Astra and BNT as well as homologous Astra and BNT regimes and also found superior humoral and cellular immune responses in the heterologous regimes compared to homologous Astra, and comparable or slightly superior immune responses when compared to homologous BNT vaccination.^3–6^ The body of research covering the effects of heterologous immunization regimes has recently been aggregated in a systematic review.^7^

*Added value of this study:* To our knowledge, this is the first study that evaluates the interdependencies of cellular and humoral immune responses following heterologous vaccination with Astra/BNT in a large group of individuals. Our data show strong correlations between humoral and cellular immune responses with the prime-boost combination Astra/BNT. The findings suggest that individuals with a robust initial response developed strong humoral and cellular immune responses after booster immunization.

*Implications of all the available evidence:* Our study and the available data suggest that due to its superior capacity to elicit a humoral and cellular immune response, mRNA-based vaccines such as BNT should be chosen for booster vaccination rather than Astra. This seems to be particularly important in individuals whose immune response was poor after initial vaccination with Astra. We demonstrate here an association between humoral and cellular immune responses following vaccination. Our findings suggest that distinct differences between common COVID-19 vaccination regimes should be taken into account in population-based vaccine programs. The present data indicate that a poor humoral immune response is unlikely to be mitigated by a strong cellular immune response.

## Introduction

COVID-19 caused by SARS-CoV-2 was declared a pandemic disease by the WHO in March 2020 and has since resulted in more than five million casualties worldwide.^8–11^ The virus enters macrophages, type II pneumocytes, pericytes and muscle cells by utilizing the angiotensin-converting enzyme 2.^12^ As a reaction, innate and acquired immune responses are mounted, including the production of antibodies and specific T cells.^13,14^ Symptoms develop approximately five to six days after infection.^15,16^

During SARS-CoV-2 infection, some individuals appear to be at higher risk for severe or even life-threatening disease. Higher age, male sex and severe preexisting health conditions are the most important determinants for such outcomes.^17^ In some patients, long-lasting symptoms occur, sometimes even after initially mild or asymptomatic disease in young and previously healthy individuals. Recently, the WHO defined “Long-COVID” as a disease course with symptoms such as fatigue, shortness of breath and cognitive dysfunction that continue for more than two months after a SARS-CoV-2 infection.^18^ Thus, it is imperative to protect the population from infection.

While SARS-CoV-2 expresses four major structural proteins (spike (S), membrane (M), envelope (E) and nucleocapsid (N) proteins), vaccine production has focused on the spike protein because of its immunogenicity and its importance for the induction of neutralising antibodies.^19–21^ Subjects inoculated with the current EMA-authorized vaccines including ChAdOx1-S (Vaxzevria, AstraZeneca, Oxford, UK), Janssen COVID-19 vaccine (Ad26.COV2.S, Janssen Vaccines in Leiden, Netherlands as a subsidiary of the American company Johnson & Johnson), BNT162b2 (Comirnaty, BioNTech, Mainz, Germany) and mRNA-1273 (Spikevax, Moderna, Cambridge, United States) develop significant levels of antibodies against the spike protein.^22–26^

Vaccines that immunize against SARS-CoV-2 epitopes provide efficient protection from the virus. Overall, levels of anti-spike RBD-IgG and neutralizing antibodies (which correlate strongly) appear to be good measures of vaccine efficacy.^27–29^ Recent publications report a vaccine efficiency of 80% against asymptomatic infections with the Alpha variant (B.1.1.7), with anti-spike-RBD-IgG titers of 506 BAU/ml or 90% efficacy for titers above 775 BAU/ml.^29,30^ Similar studies for the Delta (B.1.617.2) or Omicron (B.1.1.529) variants are expected to be published very soon.^31^ It must be assumed that higher antibody titers are necessary to prevent infections.

We are still in the process of determining optimal dosing intervals and combinations of the currently available vaccines. The ongoing debate about waning IgG titers only months after vaccination in the face of new SARS-CoV-2 variants underscores the need for a better understanding of different vaccine regimes.

When reports began to surface on cerebral venous thrombosis following Astra (ChAdOx1-S) vaccinations, particularly in younger individuals, many European governments, including Germany, restricted its use to individuals over a certain age. In Germany, this age was set at 60 years and older. This left some individuals who had been initially vaccinated with Astra insufficiently protected. As a consequence, a heterologous Astra/BNT (BNT162b2 immunization was proposed, even though at that time there was a lack of data on immunogenicity and safety.^32–35^ In the meantime, however, considerably more data have become available and these indicate that heterologous immunization with Astra/BNT is safe and at least equally effective to the homologous BNT/BNT regimen.^1–6^ Nevertheless, particularly cellular immune responses as well as individual titer developments after vaccination have not been assessed in larger cohorts. Due to the lack of comprehensive data on SARS-CoV-2-specific humoral (e.g. IgG titers against spike protein) and cellular immune responses (e.g. spike protein-directed T cell responses) on the individual level, little is known about the relationship between these two branches of the adaptive immune system.

There are associations between the level of anti-SARS-CoV-2 spike antibodies and/or the level of neutralising antibodies (nABs) and protection from symptomatic and/or asymptomatic COVID-19 following vaccination.^27–29^ However, nABs do not seem to be the sole mechanistic correlate of protection; T cells may also contribute considerably to protective immunity against SARS-CoV-2.^36^ Little is known about the relative importance of antibodies and T cells in preventing future SARS-CoV-2 infections. A first step towards clarification of this question is to assess T cells and antibodies following infection or vaccination at the individual level.

Recent research has shown declining antibody titers and waning vaccine efficacy over time.^26,37,38^ For homologous BNT, vaccine efficacy declined from 96% shortly after completion of the vaccination regime to 84% four months later.^39^ A decreasing vaccine efficacy will arguably be caused by a significant change in the immune reaction, but this is still not fully understood.

Data on the correlation between humoral and cellular vaccination responses will over time provide guidance for establishing the ideal timing for booster vaccines. The goal of this study was thus to determine and correlate IgG and T cell responses against the spike protein following heterologous Astra/BNT vaccination and compare them to individuals vaccinated with the Astra/Astra or BNT/BNT regimens, respectively.

## Materials and Methods

### Cohort

The prospective, observational COV-ADAPT cohort study was conducted at the University Medical Center Göttingen, Germany (UMG). Employees and affiliates of the UMG between the ages of 18 and 75 years who received routine first (prime) and second (booster) vaccination against COVID-19 at the UMG vaccination center were eligible for inclusion in the study unless they were currently afflicted with COVID-19 or were in domestic quarantine. The COV-ADAPT study was approved by the ethics committee of the University Medical Center Göttingen (21/5/21). Study design and study implementation were performed in accordance with the guidelines of Good Clinical Practice (ICH 1996) and the Declaration of Helsinki. The study was registered with the German Clinical Trials Register (DRKS) under (DRKS00026029).

Participants received boosters with EMA-authorized vaccines between May 14^th^ and July 14^th^, 2021. The UMG vaccination center distributed vaccines in accordance with the recommendations of the German standing committee on vaccinations (STIKO) and depending on the availability of the vaccines. Prior to study inclusion, participants had either had received an initial dose of BNT (BNT162b2; Comirnaty, BioNTech, Mainz, Germany) or Astra (ChAdOx1-S; Vaxzevria, AstraZeneca, Oxford, UK), or had had a COVID-19 infection.

After written informed consent was obtained, a questionnaire was handed out and blood samples were collected. We assessed vaccination regime, age, gender, previous COVID-19 infection or contact, medications, comorbid diseases and whether any family members were attending childcare facilities. Study subjects were only labelled “post COVID-infection”, if they had had a SARS-CoV-2 infection proven by PCR testing. At the UMG, positive SARS-CoV-2 antigen-tests are routinely verified by PCR testing and thus no study subject had a positive antigen test without subsequent confirmation by PCR testing. All other indications of a prior SARS-CoV-2 infection were summarized as “COVID contact”.

We registered and excluded all cases with immunosuppressive medication as well as those whose medication frequently has immunologically relevant side effects. The study was designed to exclude subjects with comorbidity that severely affect the immune system such as leukemia or innate immune deficiencies; however, no study participant reported such comorbid diseases. We did not exclude subjects whose concomitant diseases were expected to have no or at most moderate influence on the outcome of vaccination such as diabetes mellitus, cardiovascular disease, asthma or allergies.

### Measurement of humoral and cellular immune responses

Blood samples were collected at the UMG up to two weeks before (T1) and two weeks to three months following booster immunization (T2).

The following analyses were performed:

1. IgG antibodies directed against the receptor binding domain of the spike protein (anti-spike-RBD-IgG) via the SARS-CoV-2-IgG-II-Quant assays on the Architect i2000SR (Abbott Laboratories, Abbott Park, USA).
2. Anti-nucleocapsid-IgGs (NCP) to detect previous SARS-CoV-2 infection using the anti-SARS-CoV-2-NCP-ELISA (IgG) (Euroimmun, Lübeck, Germany) assay on the DSX Automated ELISA System (Thermo Labsystems, Chantilly, USA).
3. Neutralising antibodies (nABs) against SARS-CoV-2 through the DIA-SARS-CoV-2-nAb assay (DiaProph, Kiev, Ukraine) on the DSX Automated ELISA System (Thermo Labsystems).
4. Antibody avidity via the DIA-SARS-CoV-2-S-IgG-av avidity assay (DiaProph) on the DSX Automated ELISA System (Thermo Labsystems).
5. The cellular immune response using the SARS-CoV-2-spike-specific-IFN-γ-release assay (IGRA) (Euroimmun) on the DSX Automated ELISA System (Thermo Labsystems).

### Statistics

Statistical analyses were performed by the Scientific Core Facility for Medical Biometry and Statistical Bioinformatics (MBSB) of the UMG using the statistics software R. The significance levels were set to alpha=5% for all statistical tests.

For detailed information on methods and materials see supplementary material.

## Results

### COV-ADAPT Study Design

From May 14^th^ to July 14^th^, 2021, we recruited 417 participants between the ages of 18 and 65 years for a first blood sampling (T1) up to two weeks prior to their receiving a routine COVID-19 booster vaccination at the UMG vaccination center (see schematic in Figure 1). The average age was 35 years, with 309 female and 106 male participants. Two participants did not disclose their gender. From the 417 participants, 19 were analyzed separately for one of the following reasons: (i) a history of previously proven COVID-19 (n=8); (ii) detectable positive or border-line anti-NCP IgG antibody titers despite anamnestic negative COVID-history (n=7); (iii) insufficient information on the initial vaccine (n=1); (iv) immunosuppressive drug intake (n=3). In the latter, the immunosuppressive drugs included adalimumab (n=1), ropeginterferon alfa-2b (n=1) and apremilast (n=1).

**Figure 1:**
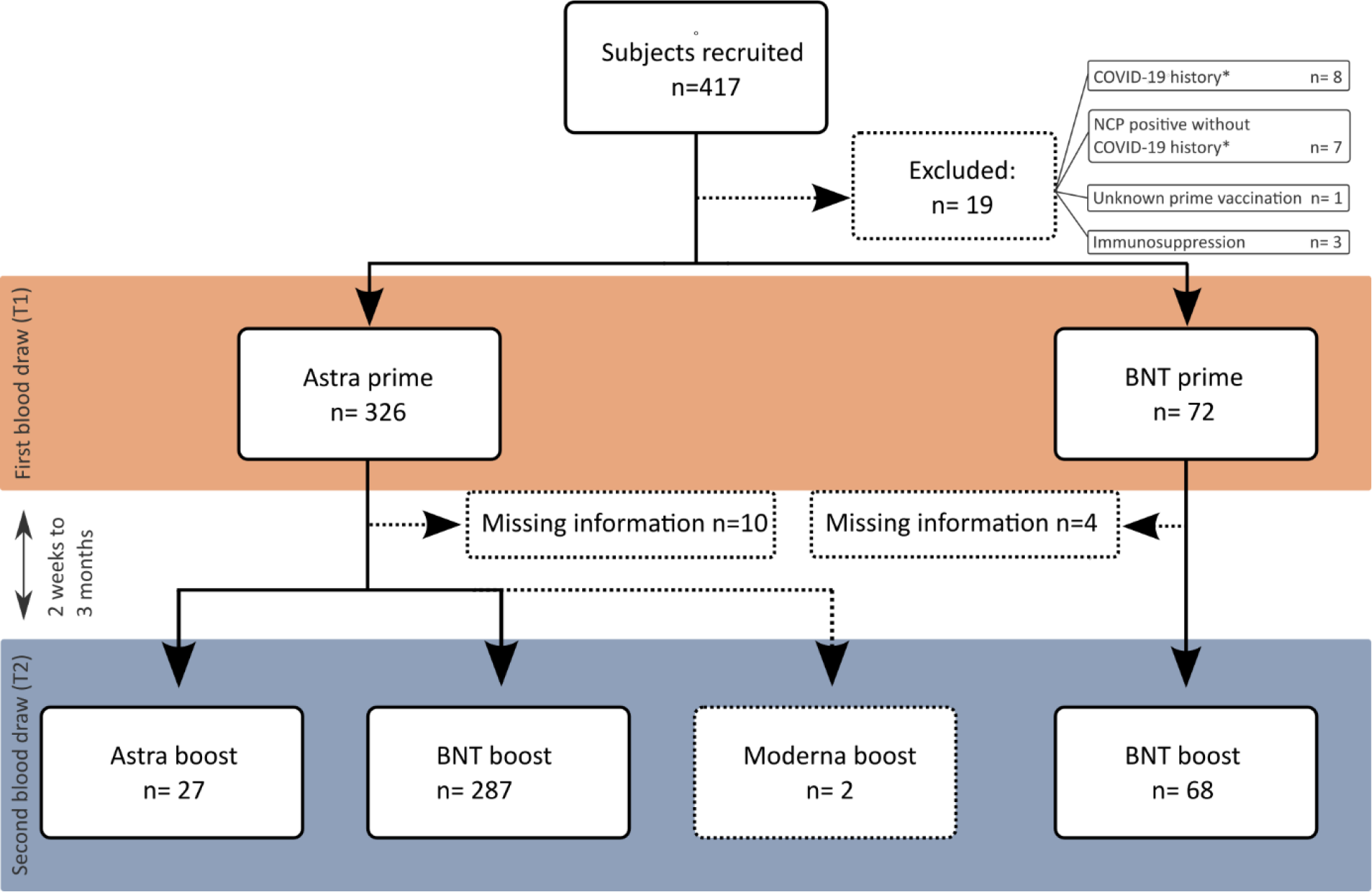
Diagram of participant recruitment and study procedure. Dashed lines indicate excluded groups. *the term “COVID-19 history” refers to PCR-confirmed SARS-CoV-2 infection.

The rest of the study participants had received either Astra as their prime vaccination (n=326 subjects) or BNT (n=72).

We asked the participants for a second blood sample to be taken 2 to 3 months after booster vaccination (T2). Of 417 invited subjects, 36 did not respond. Two of these individuals had had COVID-19 and chose not to receive a booster within the study period (n=2). Others did not show up for sampling but did provide us with information on their booster vaccination (n=20). The rest (n=14) did not respond at all, even after we tried to contact them four times. The lack of information on their booster vaccinations was noted and they were excluded from further analysis. We also excluded the 19 aforementioned participants with immune system modifications or proven contact to SARS-CoV-2 from the analysis of the second blood samples, but samples from these individuals were analyzed separately.

The remaining 384 study participants were allocated to groups according to their vaccination regimes: 27 participants had received homologous Astra/Astra, 287 had received Astra/BNT and 68 were vaccinated using homologous BNT/BNT. Two subjects had received an Astra/Moderna (mRNA-1273, Spikevax, Moderna, Cambridge, United States) combination. This group was too small in size and hence was excluded from further analysis.

The three groups (Astra/Astra, Astra/BNT, and BNT/BNT) differed somewhat in terms of participant characteristics: Based on German vaccination policies, subjects with homologous Astra vaccinations were older on average than subjects with a BNT booster (see Table 1). For further analyses, we accounted for group inhomogeneity by controlling for age and sex.

**Table 1:** Baseline characteristics and results at both time points T1 and T2. IgG = anti-spike-RBD-IgG. TC = spike-directed IFN-*γ* T cell release assay. “Total” refers to 417 recruited patients minus 8 subjects with previously proven COVID-19, 7 subjects with detectable positive or border-line anti-NCP IgG antibody titers despite anamnestic negative COVID-history and 3 subjects with immunosuppressive drug intake.

| Parameter | Total | Astra/Astra | Astra/BNT | BNT/BNT |
| --- | --- | --- | --- | --- |
| n | 399 | 27 | 287 | 68 |
| **Age** [years] |  |  |  |  |
| mean ± SD | 35 ± 13 | 57 ± 8 | 34 ± 13 | 30 ± 9 |
| **Gender** |  |  |  |  |
| male | 100 (25.1%) | 4 (14.8%) | 68 (23.7%) | 19 (27.9%) |
| female | 297 (74.4%) | 23 (85.2%) | 218 (76.0%) | 48 (70.6%) |
| unknown | 2 (0.5%) | 0 (0.0%) | 1 (0.3%) | 1 (1.5%) |
| **IgG T1** [BAU/ml] |  |  |  |  |
| mean ± SD | 94 ± 140 | 55 ± 52 | 72 ± 119 | 201 ± 194 |
| **TC T1** [mIU/ml] |  |  |  |  |
| mean ± SD | 791 ± 1803 | 1152 ± 2243 | 669 ± 1578 | 1240 ± 2509 |
| **IgG T2** [BAU/ml] |  |  |  |  |
| mean ± SD | 2370 ± 1787 | 413 ± 461 | 2387 ± 1627 | 3202 ± 2184 |
| missing | 32 | 0 | 8 | 10 |
| **TC T2** [mIU/ml] |  |  |  |  |
| mean ± SD | 4747 ± 6677 | 1680 ± 1854 | 5069 ± 6733 | 4880 ± 7570 |
| missing | 35 | 0 | 11 | 10 |
| **Avidity T1** |  |  |  |  |
| negative | 45 (11.3%) | 7 (25.9%) | 26 (9.1%) | 11 (16.2%) |
| low | 184 (46.2%) | 12 (44.4%) | 118 (41.3%) | 48 (70.6%) |
| high | 169 (42.5%) | 8 (29.6%) | 142 (49.7%) | 9 (13.2%) |
| missing | 1 | 0 | 1 | 0 |
| **Avidity T2** |  |  |  |  |
| high | 366 (100.0%) | 27 (100.0%) | 276 (100.0%) | 60 (100.0%) |
| missing | 33 | 0 | 11 | 8 |
| **Neutralization T1** |  |  |  |  |
| negative | 41 (10.3%) | 5 (18.5%) | 25 (8.7%) | 8 (11.8%) |
| positive | 357 (89.7%) | 22 (81.5%) | 261 (91.3%) | 60 (88.2%) |
| missing | 1 | 0 | 1 | 0 |
| **Neutralization T2** |  |  |  |  |
| negative | 37 (10.1%) | 0 (0.0%) | 31 (11.2%) | 6 (10.0%) |
| positive | 329 (89.9%) | 27 (100.0%) | 245 (88.8%) | 54 (90.0%) |
| missing | 33 | 0 | 11 | 8 |

### Strong primary immune response with mRNA (BNT) and high anti-spike-RBD-IgG titers after booster with BNT, independent of prime vaccination

We analyzed titers of IgG against the spike-RBD at T1 (Figure 1A) and found that individuals who had received BNT as their prime vaccination had achieved significantly higher titers, with an average of 226±279 BAU/ml (mean ± standard deviation (SD)), as compared to individuals whose prime vaccination was with Astra (70±114 BAU/ml). Samples at T2 showed that booster vaccinations significantly increased the levels of anti-spike-RBD-IgG for all included vaccine combinations (Figure 2A). However, we observed a superior anti-spike-RBD-IgG response for subjects with a BNT booster regardless of the initial vaccine, as compared to homologous Astra vaccination (Figure 2A, blue triangles). Among those with BNT booster (including the Astra/BNT and BNT/BNT group), no significant difference in anti-spike-RBD-IgG could be found at T2. Of note, anti-spike-RBD-IgG titers in individuals with a BNT booster vaccination exceeded the geometric mean of the Astra/Astra vaccinated subjects in all participants but one (Figure 2B).

**Figure 2:**
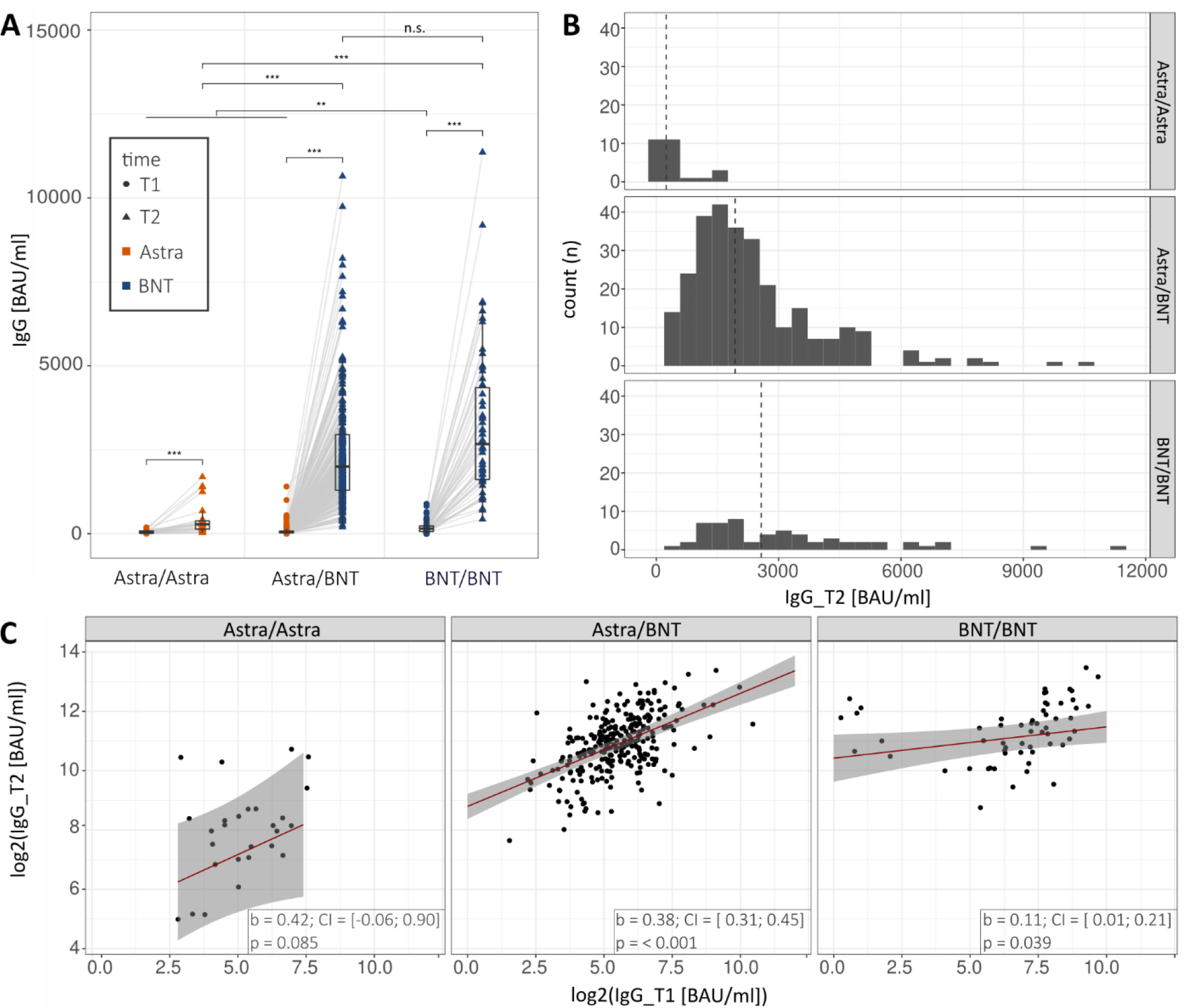
Anti-spike-RBD-IgG titers up to two weeks before (T1) and two weeks to three months after booster vaccination (T2) according to vaccination regimes. (**A**) Anti-spike-RBD-IgG (IgG) at both time points by vaccination regime. Significance asterisks indicate results from contrast tests within a linear mixed effect model for log(IgG) with vaccination regime and time and their interaction as predictors, adjusted for age and sex. The P values are adjusted for multiple testing using Holm’s procedure. *** p<0.001; ** p<0.01; * p<0.05. (**B**) Distribution (as histograms) of anti-spike-RBD-IgG measured at T2 in the different vaccination regimes (facets). The dashed lines show the geometric group means. (**C**) Regression of anti-spike-RBD-IgG at T2 (IgG_T2; second blood sample) on anti-spike-RBD-IgG at T1 (IgG_T1; first blood sample) controlling for age, sex, and time between second vaccination and T2.

A regression analsysis corrected for age, sex and elapsed time between the 2^nd^ vaccination and T2 (Figure 2C) showed a strong positive association between anti-spike-RBD-IgG titers at T1 and T2 in the Astra/BNT group (b=0.38, CI=[0.31;0.45], p<0.001) and a weaker association in the BNT/BNT group (b=0.11, CI=[0.01;0.21], p=0.039). Subjects with higher titers after prime vaccination achieved higher titers after booster vaccination. For the Astra/Astra group, a tendency towards a positive association did not reach statistical significance (b=0.42, CI=[-0.06;0.90], p=0.085).

We further analyzed whether age (Supplementary Figure 1A), time between booster and T2 (Supplementary Figure 1C), or sex (Supplementary Figure 2) impacted humoral vaccination outcomes. It was found that anti-spike-RBD-IgG titers showed a trend towards lower titers at higher ages only in the groups with a BNT booster (Supplementary Figure 1A), but not in the group that had received homologous Astra vaccine. Sex did not have a significant influence (Supplementary Figure 2). Furthermore, the magnitude of anti-spike-RBD-IgG titers changed significantly between the booster vaccination and T2 (Supplementary Figure 1B). While the antibody response tended to increase with elapsed time in the Astra/Astra group (albeit not significantly), titers declined in both groups that had received the BNT booster (significant in the Astra/BNT but not in the BNT/BNT group).

### Robust spike-directed T cell response after mRNA (BNT) booster but not after homologous Astra vaccination

Next, we measured the spike-directed IFN-γ T cell response in our study groups. There was no significant difference between persons whose prime vaccination was with either Astra or BNT at T1, with averages of 707±1631 mIU/ml and 1277±2514 mIU/ml (Figure 3A, dots). In contrast to anti-spike-RBD-IgG titers, the distribution was not strikingly different as medians partially overlapped (Figure 3B). Remarkably, there was also no statistically significant booster effect between T1 and T2 in subjects homologously vaccinated with Astra. In contrast, boostering with BNT yielded a significantly increased IFN-γ T cell response at T2, regardless of the prime vaccination (Figure 3A, blue triangles). Similar to the findings with the anti-spike-RBD-IgG, there was no significant difference in the T cell response between the Astra/BNT and the BNT/BNT group.

**Figure 3:**
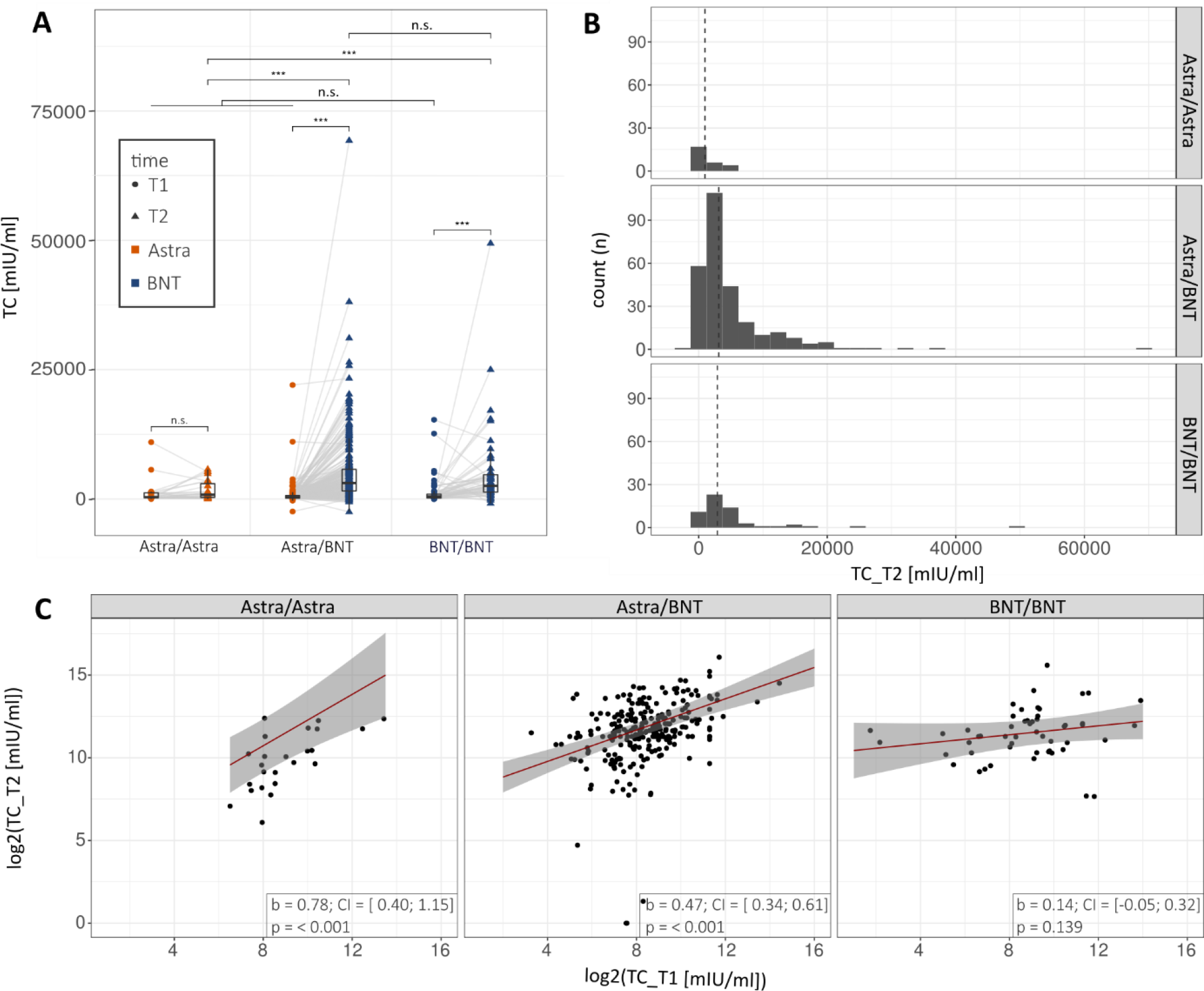
T cell responses as measured with spike-directed IFN-γ T cell response assay (IGRA) up to 2 weeks before (T1) and 2 weeks to 3 months after booster vaccination (T2) according to vaccination regimes, as indicated. (**A**) Spike-directed IFN-γ T cell responses as measured with IGRA at both blood sampling by vaccination regime. Significance asterisks indicate results from contrast tests within a linear mixed effect model for log(TC) with vaccination regime and time and their interaction as predictors and additionally adjusted for age and sex. The P values are adjusted for multiple testing using Holm’s procedure. *** p<0.001; ** p<0.01; * p<0.05. (**B**) Distribution (as histograms) of IGRA results measured at T2 in the different vaccination regimes (facets). The dashed lines show the geometric group means. (**C**) Regression of TC_T2 (Anti-spike IGRA after the second blood sample) on TC_T1 (Anti-spike IGRA after the first blood draw) controlling for age, sex, and time between second vaccination and T2.

Regression analysis between T cell responses at T1 and T2 (Figure 3C) showed significant linear associations particularly for the Astra/Astra group (b=0.78, CI=[0.40;1.15], p<0.001). For the Astra/BNT group there was also a positive association (b=0.47, CI=[0.34;0.61], p<0.001), whereas for the BNT/BNT group no association could be detected (b=0.14, CI=[-0.05;0.32], p=0.139.

When examining the parameters age (Supplementary Figure 1B), time between booster and T2 (Supplementary Figure 1D), and sex (Supplementary Figure 2) with respect to IFN-γ T cell response, it was weakly (not statistically significantly) increased with age only in the Astra/Astra group. Our data points towards a slight reduction in spike-directed IFN-γ T cell activity depending on the elapsed time between booster vaccination and T2 for the Astra/BNT group. This effect could not be observed for BNT/BNT or Astra/Astra vaccination individuals (Supplementary Figure 1D).

### Associations between humoral and cellular immune responses in primary and booster vaccination

To assess the association between anti-spike-RBD-IgG titers and spike-directed IFN-γ T cell responses (Figure 4), separate regression analyses for T1 (Figure 4A and B) and T2 (Figure 4C and D) were performed. The analysis was conducted for all study participants combined (Figures 4A and C) as well as for the different vaccination regimes (Figures 4B and D). For the study group as a whole, there was a clear positive association between anti-spike-RBD-IgG titers and spike-directed IFN-γ T cell responses for both T1 and T2 (b=0.21, CI=[0.10;0.32], p<0.001 for T1 and b=0.35, CI=[0.17;0.53], p<0.001 for T2). However, when considering vaccination groups separately, significant correlations were only found at T1 in the Astra/Astra and the Astra/BNT groups, whereas no association was seen in the BNT/BNT group at T1. At T2, none of the vaccine regimes yielded a significant correlation, although in the Astra/BNT group a trend was detectable (b=0.26, CI=[0.00;0.53], p=0.054).

**Figure 4:**
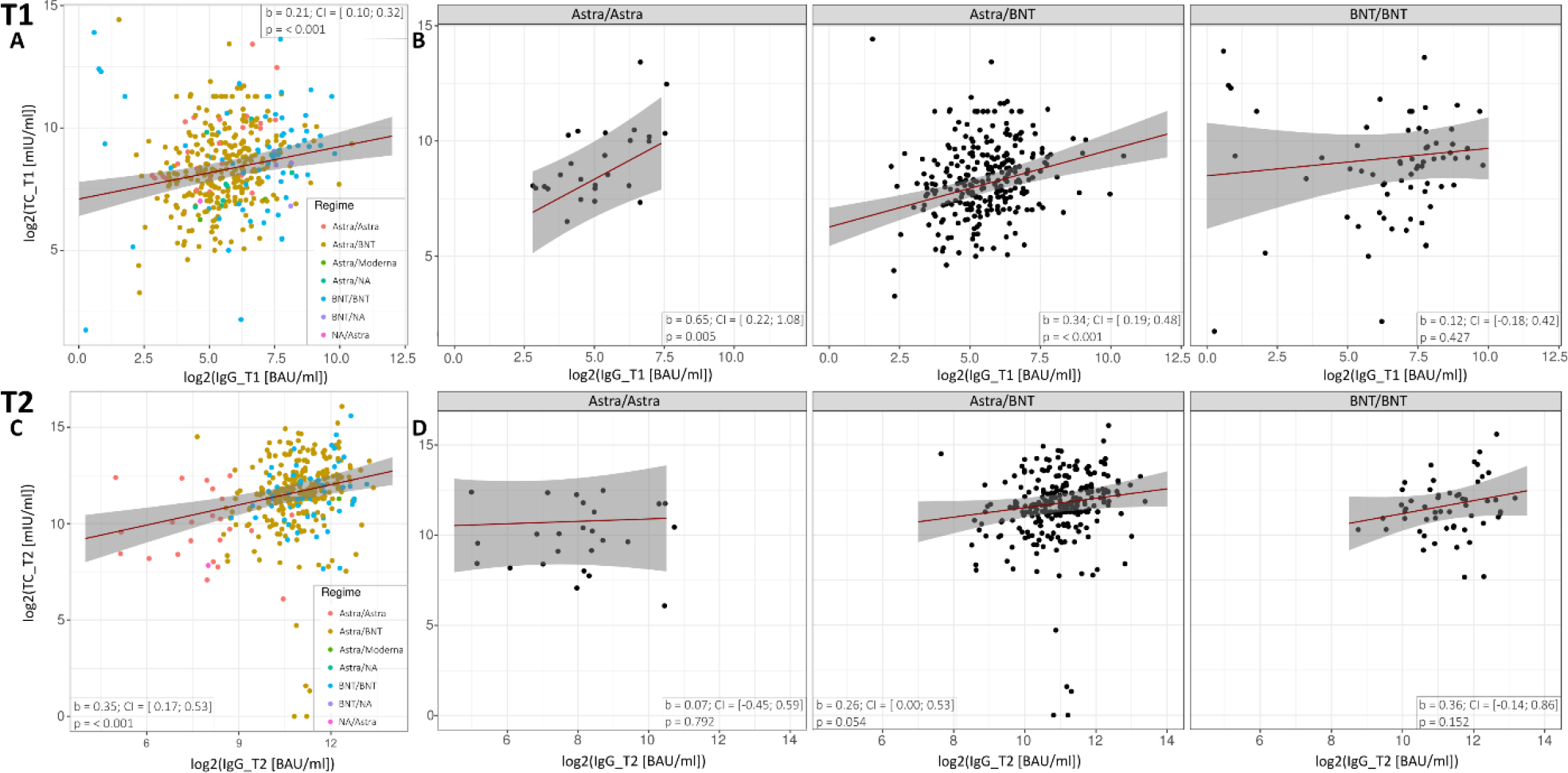
Regression of the spike-directed IFN-γ T cell responses (TC) and anti-spike-RBD-IgG (IgG) at the first (T1) and the second blood sample (T2). (**A**) Regression at T1 of log2(TC_T1) on log2(IgG_T1) controlling for age, sex, and time between first vaccination and T1 for all study participants and (**B**) for the respective subgroups. (**C**) Regression at T2 of log2(TC_T2) on log2(IgG_T2) controlling for age, sex, and time between second vaccination and T2 in all study participants and (**D**) for the respective subgroups.

An additional analysis investigated the correlation between the spike-directed T cell IFN-*γ* responses at T1 and anti-spike-RBD-IgG at T2 (Figure 5). Significant correlations were found for the Astra/BNT and BNT/BNT group, e.g. good T cell responses after T1 were correlated with a good antibody response at T2 (b=0.14, CI=[0.06;0.21], p<0.001 for Astra/BNT and b=0.11, CI=[0.01;0.21], p=0.026 for BNT/BNT).

**Figure 5:**
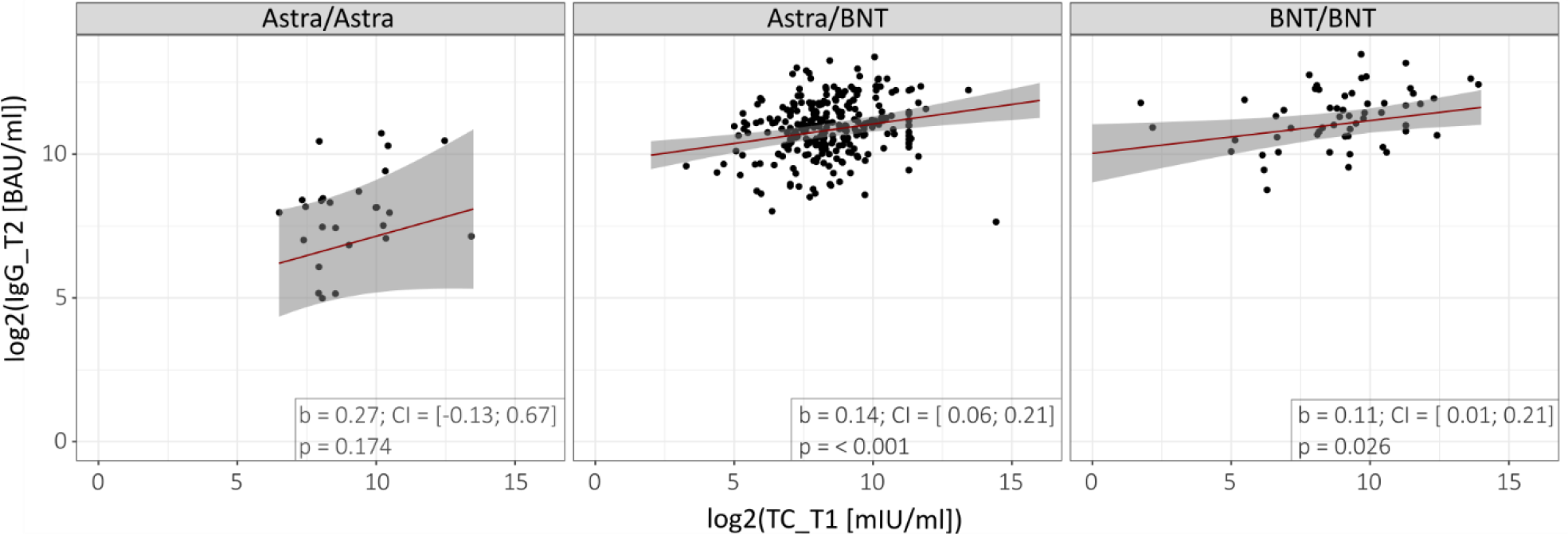
Association of the spike-directed IFN-γ T cell responses (TC) at the first blood sample (T1) and anti-spike-RBD-IgG (IgG) at the second blood sample (T2). Regression of log2(IgG_T2) on log2(TC_T1) controlling for age, sex, and time between first vaccination for the respective subgroups.

### Neutralizing antibodies and increasing antibody avidity in all three regimens

The neutralization index (NI) as a parameter for the development of neutralizing antibodies against SARS-CoV-2 (Figure 6a) as well as the relative avidity index (RAI) (Figure 6b) were also determined for both time points T1 and T2. Both qualitative parameters complement the quantitative determination of the antibody titers In all three groups (Astra/Astra, Astra/BNT and BNT/BNT), there were measurable titers of neutralizing antibodies already at T1. We found a significant increase in the NI from T1 to T2, most pronounced in both the Astra/Astra and the Astra/BNT groups, and more moderate in the BNT/BNT group. The moderate increase in the BNT/BNT group can be attributed to the already high level of neutralization indices at T1 (Figure 6).

**Figure 6:**
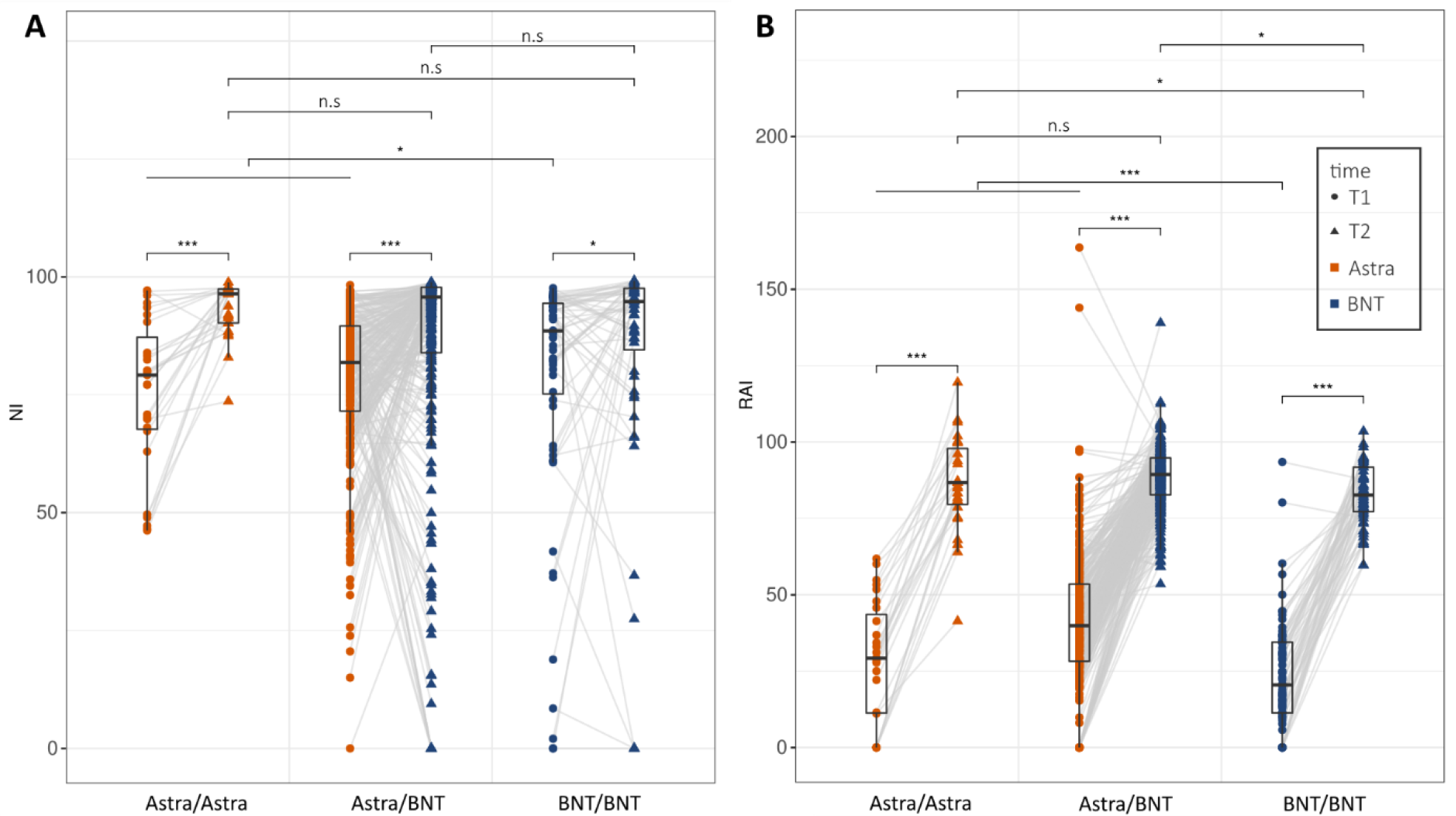
Neutralization index (NI) (**A**) and relative avidity index (RAI) (**B**) at both time points depicted according to vaccination regimens. Significance asterisks indicate results from contrast tests within a linear mixed effect model for RAI with vaccination regime and time and their interaction as predictors and additionally adjusted for age and sex. The P values are adjusted for multiple testing using Holm’s procedure. *** p<0.001; ** p<0.01; * p<0.05.

Interestingly, in both BNT-boostered groups, a small number of participants (31/276 for the Astra/BNT group and 6/60 for the BNT/BNT group) presented with no neutralizing antibodies at T2, even though the majority of these participants had had measurable neutralizing antibodies at T1. Only three subjects showed neither neutralizing antibodies at T1 nor at T2, all of whom had received homologous BNT. Of note, the lack of neutralizing antibodies did not correspond to a lack of anti-spike-RBD-IgG, as these participants did have above average anti-spike-RBD-IgG titers.

Avidity, a measure for the binding strength of a multivalent bond between antigen and antibody, is often used as a parameter of the quality of an antibody response. In our study, persons who were prime vaccinated with Astra reached a significantly higher avidity index at T1 when compared to BNT (Figure 6B). All groups showed a significant increase in avidity at T2. Interestingly, the BNT/BNT group also reached the lowest avidity at T2, which was significantly lower than for both groups with prime vaccination with Astra. We thus found that the ratio between antibody titers and antibody avidity is different between the vaccination schemes. The reason for this could either be that (i) Astra yields fewer but more avid antibodies or (ii) our avidity assay is insufficiently quantitative.

## Discussion

COV-ADAPT is an observational cohort study providing real-world data on heterologous vaccination with Astra/BNT compared to homologous Astra/Astra and BNT/BNT-vaccinations in a large cohort of 417 health care workers. As the focus of this study was the evolution of the immune response after prime and booster vaccination, baseline values of participants before both vaccines were not included. We correlated immune responses on an individual level after prime vaccination with the responses after booster to evaluate the predictability of the quality of the immune response. We were able to demonstrate that humoral and cellular immune responses correlate with one another, suggesting that a poor humoral immune response is unlikely to be ameliorated by a strong cellular response.

In our study, we found a superior effectiveness in regard to immune stimulation for the mRNA-based vaccines (i.e. BNT), either in a homologous BNT/BNT vaccine regime or as a heterologous combination with Astra (Astra/BNT). This superior effectiveness was observed both in terms of the humoral immune response (anti-spike-RBD-IgG titers) as well as the cellular component, i.e. the spike-directed IFN-γ T cell responses. Our findings corroborate with previous studies which also demonstrated that the IgG antibody response against the spike protein in the heterologous Astra/BNT regime is comparable to the homologous BNT/BNT and superior to the homologous Astra/Astra regime.^2–6^ Furthermore, these studies showed that heterologous immunization with Astra/BNT yields a tolerable and manageable reactogenicity compared to homologous Astra and BNT vaccinations.^1–4,6^

The combination of vector- and mRNA-based vaccines was initially thought to provide the benefits of both vaccination techniques. The hope was that this combination would yield the strong IgG responses known from mRNA vaccines as well as enhanced T cell responses.^40^ Both responses appear to play an important role in vaccine-induced protection against SARS-CoV-2 infection, particularly in the early phase after vaccination.^21,36^ CD8^+^ cell responses in particular were expected to be increased through heterologous vaccination with Astra/BNT, as based on animal models and pathophysiological considerations.^40,41^ Several studies in smaller subpopulations appeared to support this hypothesis.^2,3,42^ However, in our large data set, we were not able to confirm a general superiority of the Astra/BNT regime compared to homologous mRNA vaccination (BNT/BNT) in terms of T cell stimulation. At T2, booster vaccination with BNT significantly increased spike-directed IFN-γ release of T cells in individuals from the Astra/BNT regime as well as the BNT/BNT regime, with no statistically relevant difference between these two groups.

Booster vaccination with Astra had no discernible influence on IFN-γ release by T cells, suggesting that booster immunization in this subgroup is ineffective for increasing T cell-mediated immunity. This is in line with recent findings by Hillus et al.^6^, who analyzed spike-specific IFN-*γ*-release of T cells in a small cohort and found that Astra booster vaccination did not further increase the T cell response in individuals prime vaccinated with Astra. Of note, Schmidt et al.^3^ showed in a small study that all three vaccination regimes (Astra/Astra, Astra/BNT, BNT/BNT) induced polyfunctional T cells, which slightly differed in the induced CD4^+^ and CD8^+^ T cell subpopulations, distinguishable by their particular cytokine profiles. We did not account for different T cell subpopulations and can hence not exclude subtle functional advantages of a heterologous vaccination.

We designed the COV-ADAPT study to assess correlations between immune responses from the first to the second vaccination. We found that the humoral (anti-spike-RBD-IgG) response at T1 was positively correlated with the response at T2 for both BNT-boostered groups, with a stronger effect measured in the Astra/BNT group. The correlation within the Astra/Astra group showed a similar effect but did not reach statistical significance. This may be attributed to the small group size (n=27). Thus, a strong or a weak initial humoral response appears to be predictive for the development of a high or a low titer of protective anti-spike-RBD-IgG at T2, respectively.

Additionally, we found a strong and significant positive correlation between IFN-*γ* release by T cells at T1 and T2 for the Astra/Astra and the Astra/BNT groups. Thus, participants who started off with a strong response also showed strong responses at T2, while weak responses remained weak at T2. The positive correlation was not associated with an increased T cell response from T1 to T2 (Figure 3A) for the Astra/Astra group. This may be attributed to the fact that T cell activity depended heavily on the reaction to the prime vaccine and could not be further increased by the Astra booster. In contrast, in the Astra/BNT group T cell responses generally increased from T1 to T2, indicating a BNT-induced booster effect.

On an aggregate level, humoral and cellular responses correlated with each other at T1 and T2, respectively. When looking at the particular vaccine regimes, a significant association between these responses could only be detected for the Astra/Astra and the Astra/BNT group at T1. The loss of significance for all vaccination regimes at T2 could be attributed to a stronger augmentation auf anti-spike-RBD-IgG titers compared to the T cell responses. One may speculate that antibody titers have a higher capacity to increase as compared to the cellular compartment of the immune system, resulting in a loss of linear association.

An additional analysis investigating the correlation between IFN-*γ* response at T1 and anti-spike-RBD-IgG at T2 showed a significant correlation for the Astra/BNT and BNT/BNT groups, suggesting that good T cell responses after T1 corresponded with good anti-spike-RBD-IgG responses at T2 (Figure 5). This is in line with the notion that functional T helper cells as well as memory T cells are fundamental for launching a successful immune response.^43^ Thus, an effective spike-directed T cell activation at T1 leads to increased anti-spike-RBD titers at re-exposure to the antigen.

While quantitative immune responses did not differ between males and females, subjects of higher age and male sex showed lower avidity after the prime vaccination (Supplementary Table 2). Such general sex-dependent differences in immune responses might contribute to increased COVID-19 severity in men and older individuals, as has been observed in previous studies.^17^ However, due to the semi-quantitative nature of the test used here, all subjects reached the maximum test category “high avidity” at T2 (Figure 6). We therefore did not find a correlation between vaccination regimes and/or specific participant characteristics.

Most individuals who received BNT as a booster developed a significant immune response, which was, by all quantitative parameters measured in this study, highly superior to immunity induced by Astra/Astra. Unexpectedly, there were no significant differences between the three vaccination regimens in regard to the neutralization indices at T1 and T2. A possible explanation is that the relative neutralization capacity had already been high (90%) after prime vaccination. However, in contrast to the Astra/Astra group, some individuals presented with a lack of neutralizing antibodies at T2 following booster vaccination with BNT. Additionally, there were hints at a more short-lived immune response after BNT booster even within the relatively short observation time, as compared to the Astra/Astra vaccination (Supplementary Figure 1). This would be in line with other studies showing a faster decline in humoral responses following mRNA vaccinations compared to vector-based vaccines.^26,31^ It should be taken into account that the Astra carrier virus DNA remains in the organism in a transcriptionally active form for months, while maintaining activated CD8-T cells.^44^ In contrast, persistence of mRNA in the organism is short (at least in mice^45^), with a maximum of 10 days. It is conceivable that prolonged spike protein exposure in vector-based vaccines enables the organism to increase the quality of the immune response over a longer time.

In conclusion, our study suggests that an mRNA-based vaccine such as BNT should generally be chosen for booster vaccination rather than Astra due to its superiority in the elicited humoral and cellular immune response. This appears particularly important for those individuals with a poor immune response after prime vaccination with Astra.

Our findings revealed an association between humoral and cellular immune responses following vaccination, and that a poor humoral immune response is unlikely to be mitigated by a strong cellular immune response. The distinct differences between vaccination regimes, as demonstrated in this study, should be taken into account for population-based vaccine programs.

## Data Availability

Data and materials of this study are available from the authors on request.

## Declarations

### Ethics approval and consent to participate

The COV-ADAPT study was approved by the ethics committee of the University Medical Center Göttingen (21/5/21). Informed consent was obtained from all participants and study design and study implementation were performed in accordance with the guidelines of Good Clinical Practice (ICH 1996) and the Declaration of Helsinki. The study was registered with the German Clinical Trials Register (DRKS) under (DRKS00026029).

### Consent for publication

All authors reviewed the final manuscript version and consented to its submission.

### Availability of data and materials

Data and materials of this study are available from the authors on request.

## Acknowledgments

We would like to thank Carsten Watzl for his scientific input and thorough review of the manuscript.

## Contribution of each author

LE, MS, MH and AZ designed the study with input from MPS, UG, HSB and JS. LE, MH, MS, LM, AB, TMH, AA, AE, JSH, KZ, MS and AS recruited patients and performed experiments. MH and LM collected data, MH and AL extracted and compiled data; all authors discussed data; MH, LE and MS drafted the manuscript; all authors jointly discussed, reviewed and amended the manuscript. Data verification was performed by MH and LM.

---

STROBE Statement—Checklist of items that should be included in reports of ***cohort studies***

|  | Item No | Recommendation |  |
| --- | --- | --- | --- |
| Title and abstract | 1 | (a) Indicate the study’s design with a commonly used term in the title or the abstract | ✓ |
|  |  | (b) Provide in the abstract an informative and balanced summary of what was done and what was found | ✓ |
| Introduction |  |  |  |
| Background/rationale | 2 | Explain the scientific background and rationale for the investigation being reported | ✓ |
| Objectives | 3 | State specific objectives, including any prespecified hypotheses | ✓ |
| Methods |  |  |  |
| Study design | 4 | Present key elements of study design early in the paper | ✓ |
| Setting | 5 | Describe the setting, locations, and relevant dates, including periods of recruitment, exposure, follow-up, and data collection | ✓ |
| Participants | 6 | (a) Give the eligibility criteria, and the sources and methods of selection of participants. Describe methods of follow-up | ✓ |
|  |  | (b) For matched studies, give matching criteria and number of exposed and unexposed | ✓ |
| Variables | 7 | Clearly define all outcomes, exposures, predictors, potential confounders, and effect modifiers. Give diagnostic criteria, if applicable | ✓ |
| Data sources/ measurement | 8* | For each variable of interest, give sources of data and details of methods of assessment (measurement). Describe comparability of assessment methods if there is more than one group | ✓ |
| Bias | 9 | Describe any efforts to address potential sources of bias | ✓ |
| Study size | 10 | Explain how the study size was arrived at | - |
| Quantitative variables | 11 | Explain how quantitative variables were handled in the analyses. If applicable, describe which groupings were chosen and why | ✓ |
| Statistical methods | 12 | (a) Describe all statistical methods, including those used to control for confounding | ✓ |
|  |  | (b) Describe any methods used to examine subgroups and interactions | ✓ |
|  |  | (c) Explain how missing data were addressed | ✓ |
|  |  | (d) If applicable, explain how loss to follow-up was addressed | ✓ |
|  |  | (e) Describe any sensitivity analyses | ✓ |
| Results |  |  |  |
| Participants | 13* | (a) Report numbers of individuals at each stage of study—eg numbers potentially eligible, examined for eligibility, confirmed eligible, included in the study, completing follow-up, and analysed | ✓ |
|  |  | (b) Give reasons for non-participation at each stage | ✓ |
|  |  | (c) Consider use of a flow diagram | ✓ |
| Descriptive data | 14* | (a) Give characteristics of study participants (eg demographic, clinical, social) and information on exposures and potential confounders | ✓ |
|  |  | (b) Indicate number of participants with missing data for each variable of interest | ✓ |
|  |  | (c) Summarise follow-up time (eg, average and total amount) | ✓ |
| Outcome data | 15* | Report numbers of outcome events or summary measures over time | ✓ |
| Main results | 16 | (a) Give unadjusted estimates and, if applicable, confounder-adjusted estimates and their precision (eg, 95% confidence interval). Make clear which confounders were adjusted for and why they were included | ✓ |
|  |  | (b) Report category boundaries when continuous variables were categorized | ✓ |
|  |  | (c) If relevant, consider translating estimates of relative risk into absolute risk for a meaningful time period | n.r. |
| Other analyses | 17 | Report other analyses done—eg analyses of subgroups and interactions, and sensitivity analyses | ✓ |
| **Discussion** |  |  |  |
| Key results | 18 | Summarise key results with reference to study objectives | ✓ |
| Limitations | 19 | Discuss limitations of the study, taking into account sources of potential bias or imprecision. Discuss both direction and magnitude of any potential bias | ✓ |
| Interpretation | 20 | Give a cautious overall interpretation of results considering objectives, limitations, multiplicity of analyses, results from similar studies, and other relevant evidence | ✓ |
| Generalisability | 21 | Discuss the generalisability (external validity) of the study results | ✓ |
| **Other information** |  |  |  |
| Funding | 22 | Give the source of funding and the role of the funders for the present study and, if applicable, for the original study on which the present article is based | ✓ |

\*Give information separately for exposed and unexposed groups.
**Note:** An Explanation and Elaboration article discusses each checklist item and gives methodological background and published examples of transparent reporting. The STROBE checklist is best used in conjunction with this article (freely available on the Web sites of PLoS Medicine at http://www.plosmedicine.org/, Annals of Internal Medicine at http://www.annals.org/, and Epidemiology at http://www.epidem.com/). Information on the STROBE Initiative is available at http://www.strobe-statement.org.

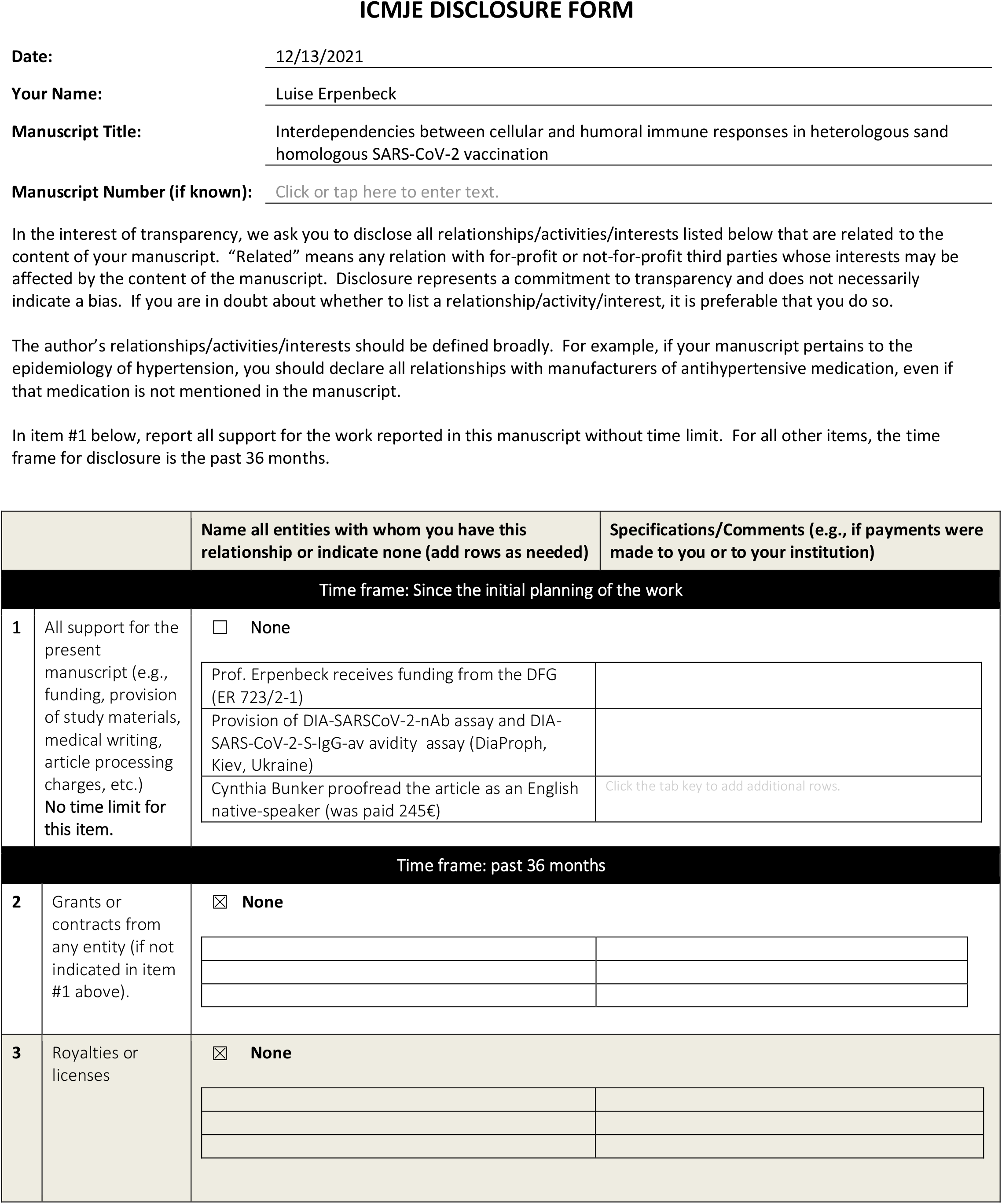
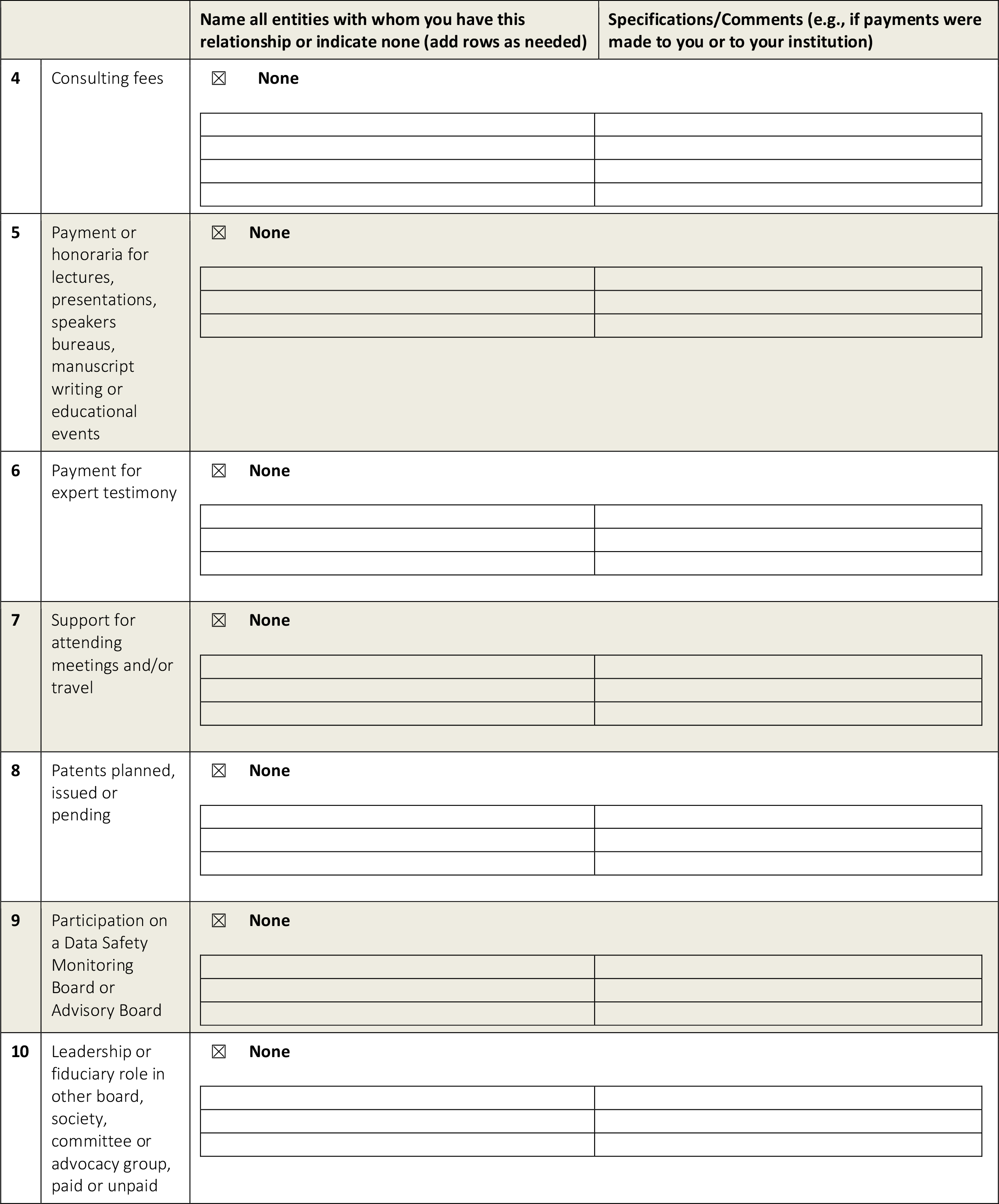
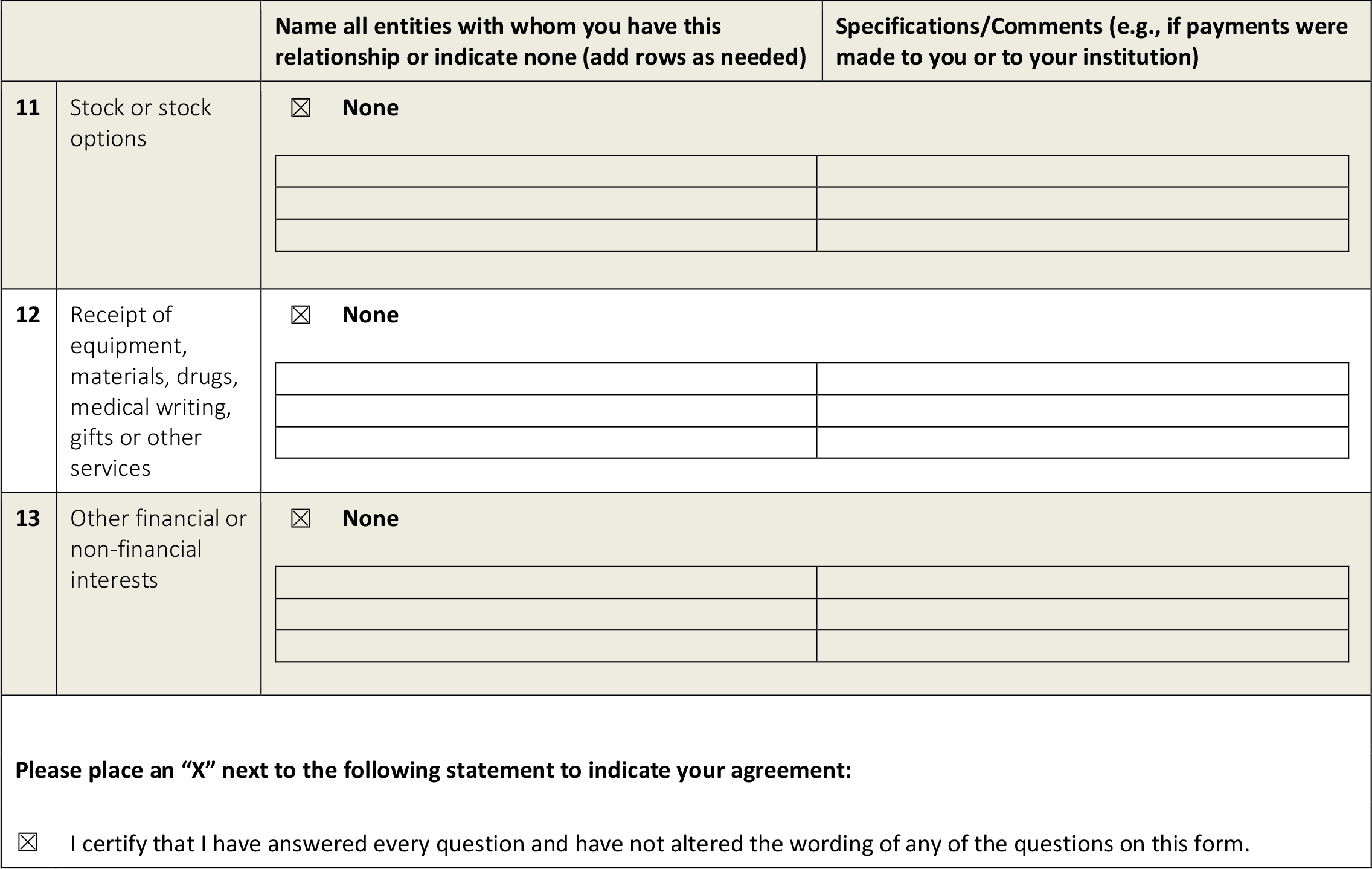

