## Supplementary Files for "Interdependencies between cellular and humoral immune responses in heterologous and homologous SARS-CoV-2 vaccination"

### Supplements

### (A) Supplementary Figures

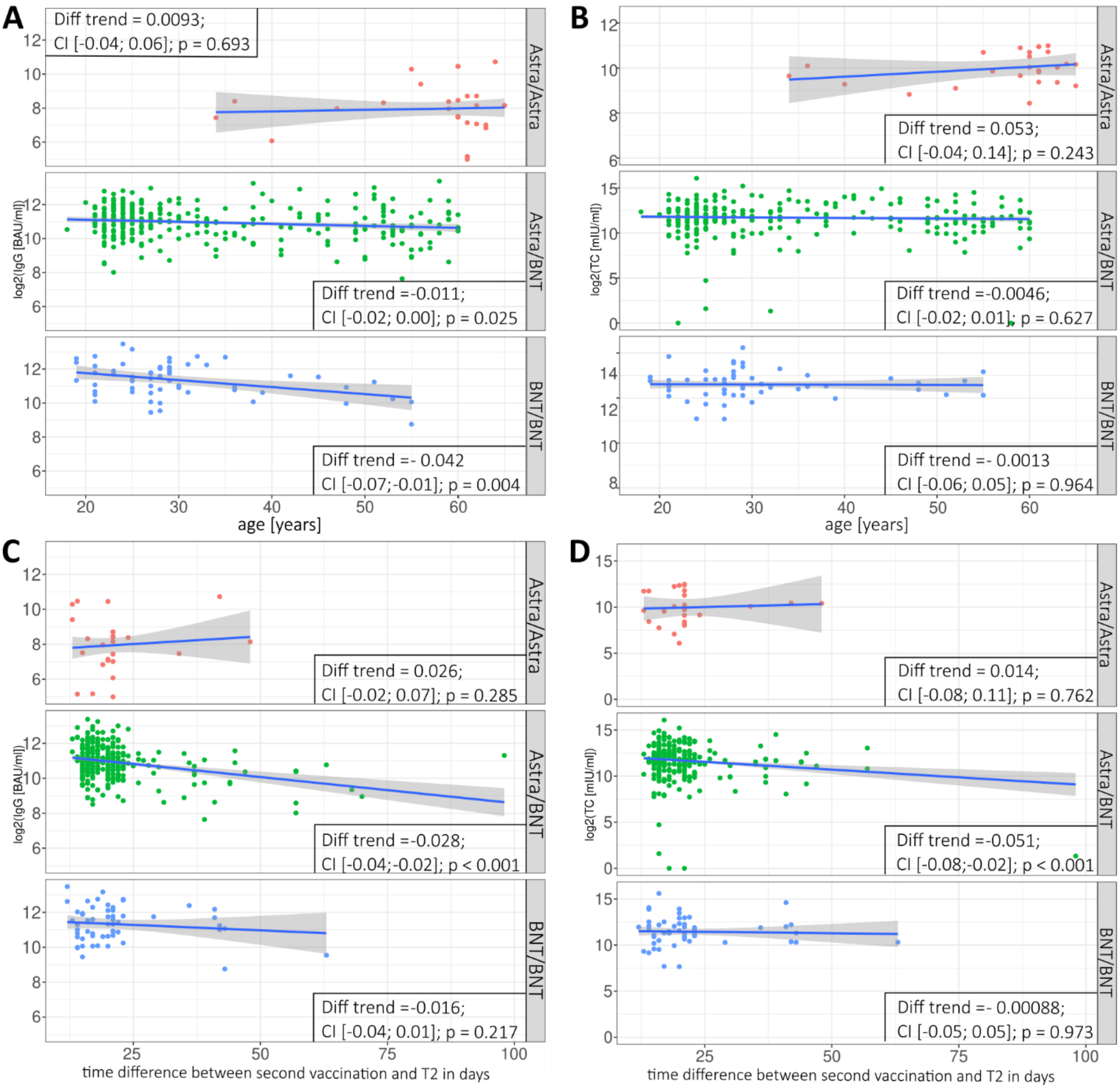


***Supplementary Figure 1:*** *Correlation between anti-spike-RBD-IgG (IgG) (left; A and C) and spike-directed IFN-γ T cell response (TC) (right; B and D) with age (top; A and B) and with the time period between booster vaccination and T2 (bottom; C and D). Study groups (Astra/Astra, Astra/BNT, BNT/BNT) as indicated. In boxes modelled trends of IgG and TC at T2 dependent on the time (in days) between the second vaccination and T2. IgG and TC at T2.*

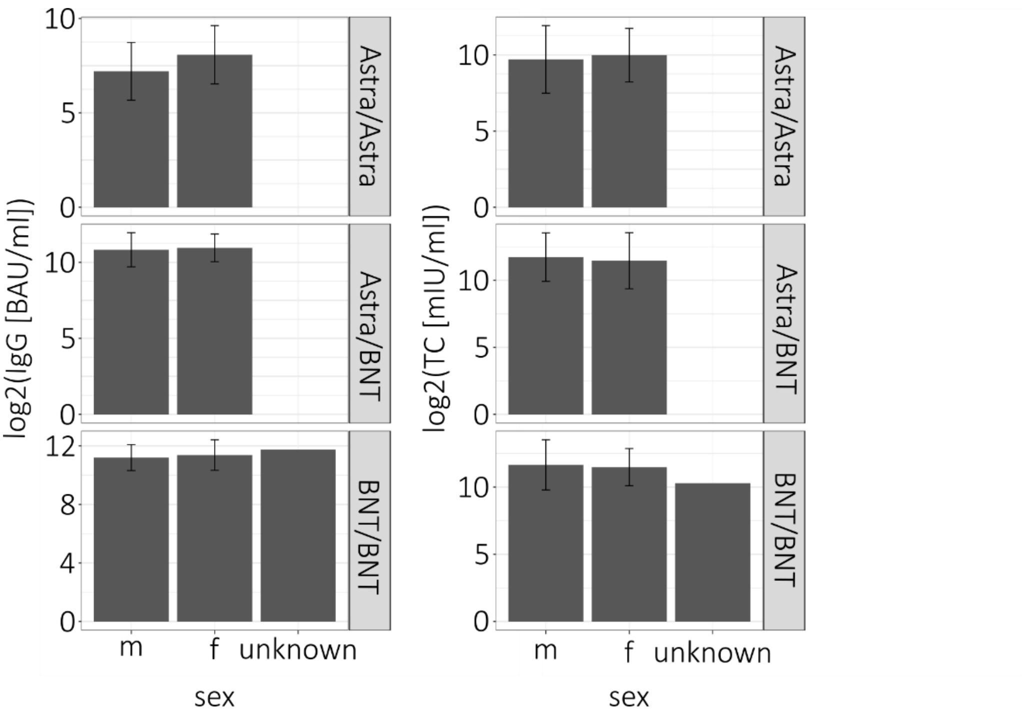


***Supplementary Figure 2:*** *Anti-spike-RBD-IgG titers (IgG) and T cell responses (TC) by sex grouped by regimen. Left: TC, right: IgG. The bars visualize the mean, the error bars the standard deviation.*

### (B) Supplementary Tables

***Supplementary Table 1:*** *Neutralization [NI]: Model coefficients from a linear mixed effect model for NI with vaccination regime and time and their interaction as predictors and additionally adjusted for age and sex. For each term in the model, the estimated effect is given with 95% confidence interval.*

| Term | Estimate | ci |
|---|---|---|
| **(Intercept)** | 1.1 | [ 0.58; 1.69] |
| **RegimeAstra_BNT** | 0.32 | [-0.12; 0.75] |
| **RegimeBNT_BNT** | 0.68 | [ 0.17; 1.18] |
| **TimeT2** | 1.3 | [ 0.77; 1.85] |
| **Age** | 0.0097 | [ 0.00; 0.02] |
| **Gender (female)** | -0.0043 | [-0.19; 0.18] |
| **Gender unknown** | 1.2 | [-0.30; 2.69] |
| **RegimeAstra_BNT:timeT2** | -0.34 | [-0.91; 0.23] |

| | | |
|---|---|---|
| **RegimeBNT_BNT:timeT2** | -0.73 | [-1.38; -0.08] |

***Supplementary Table 2:*** *<u>Avidity [RAI]:</u> Model coefficients from a linear mixed effect model for RAI with vaccination regime and time and their interaction as predictors, additionally adjusted for age and sex. For each term in the model, the estimated effect is given with 95% confidence interval.*

| Term | Estimate | ci |
|---|---|---|
| **(Intercept)** | 43 | [ 34.29; 52.31] |
| **RegimeAstra_BNT** | 5.7 | [ -1.39; 12.82] |
| **RegimeBNT_BNT** | -12 | [-20.11; -3.74] |
| **TimeT2** | 58 | [ 49.35; 66.57] |
| **Age** | -0.3 | [ -0.41; -0.19] |
| **Gender (female)** | 2.8 | [ -0.19; 5.81] |
| **Gender unknown** | 3.7 | [-20.79; 28.19] |
| **RegimeAstra_BNT:timeT2** | -11 | [-19.68; -1.63] |
| **RegimeBNT_BNT:timeT2** | 0.61 | [ -9.75; 10.98] |

### (C) Supplementary Material: Materials and Methods

Blood collection

Blood samples were collected in the University Medical Center Göttingen less than 2 weeks before (T1) and 2 weeks to 3 months following booster immunization (T2). For each blood sampling, two Sarstedt-S monovettes® of 7·5ml were taken (1x lithium heparin, 1x serum).

Measurement of anti-spike-RBD-IgG titers

To monitor antibody responses following vaccination, automated SARS-CoV-2-IgG-II-Quant assays (Abbott Laboratories, Abbott Park, USA) were used on the Architect i2000SR (Abbott Laboratories) at the central laboratory of the UMG. For this analysis, study participants´ serum was measured either immediately after collection or stored for a maximum of 7 days at -20°C before being analyzed according to the manufacturer's instructions. Values exceeding 40,000 AU/mL were diluted 1:2, and measurements above the cut-off of 50 AU/mL were considered positive according to the manufacturer´s instructions. The SARS-CoV-2-IgG-II-Quant is a chemiluminescent microparticle immunoassay (CMIA) which detects IgG antibodies, including neutralizing antibodies, in serum against the receptor binding domain (RBD) of the S1 subunit of the SARS-CoV-2 spike protein. AU/mL values were converted to binding antibody units (BAU)/mL by multiplying them with 0.142 according to the manufacturer´s instructions.

Anti-SARS-CoV-nucleocapsid (NCP) ELISA

The SARS-CoV-2-IgG-II-Quant assay does not distinguish a positive response between IgGs following SARS-CoV-2 vaccination or infection. The anti-SARS-CoV-2-NCP-ELISA (IgG) (Euroimmun, Lübeck, Germany) assay was conducted to specifically identify IgGs following SARS-CoV-2 infection, following the manufacturer's instructions and using the DSX Automated ELISA System (Thermo Labsystems).

SARS-CoV-2 spike-specific IFN-γ release assay

The SARS-CoV-2-spike-specific-IFN-γ-release-assay (IGRA) (Euroimmun) was performed according to the manufacturer's instructions to evaluate the T cell response. Briefly, three heparinized full-blood samples of 0.5 ml were processed from each participant. One sample was incubated with the SARS-CoV-2 spike S1 peptide pool, one with a mitogen (positive control), and one remained unstimulated (negative control). All three samples were incubated overnight. The supernatants were stored at -20° until analysis. Thawed samples were diluted 1:5 with dilution buffer and the automated IFN-γ ELISA was performed with the DSX Automated ELISA System (Thermo Labsystems).

The results were calculated by subtracting the IFN-γ concentration (in mIU/ml) of the blank sample tube from that of the spike-stimulated test tube. Samples for which the result exceeded 2,500 mIU/ml were re-measured using a dilution of 1:15, 1:20, 1:50, 1:100 and when necessary 1:200 with the automated IFN-γ release ELISA-testing.

Measurements of neutralizing antibodies and antibody avidity

Subsequently, each patient's frozen plasma was screened for neutralizing antibodies against SARS-CoV-2 and analyzed for their respective antibody avidity. We used the competitive DIA-SARS-CoV-2-nAb ELISA (DiaProph, Kiev, Ukraine) and the DIA-SARS-CoV-2-S-IgG-av avidity assay (DiaProph) based on the mammalian expressed RBD antigen (AA 320-524) and performed the tests fully automated on the DSX Automated ELISA System (Thermo Labsystems) following the manufacturer's instructions.

Statistics

Statistical analyses were performed by the Scientific Core Facility for Medical Biometry and Statistical Bioinformatics (MBSB) of the UMG.

All variables were summarized for the full cohort as well as for each vaccination regime using absolute and relative frequencies or mean ± standard deviation, as appropriate. Immune responses, as anti-spike-RBD-IgG and spike-directed IFN-γ T cell response) as well relative avidity index (RAI) and neutralization index (NI) were modelled using linear mixed effect models with vaccination regime and time and their interaction as predictors, and were additionally adjusted for age and sex. Anti-spike-RBD-IgG and spike-directed IFN-γ T cell responses were modelled on a log scale, NI on a reverse log scale. We reported resulting model coefficients with 95% confidence intervals. Pairwise contrast tests were conducted to compare time points within each vaccination regime as well as between the vaccination regimes at each time point. Resulting p values were adjusted for multiple testing using Holm's procedure.

The time difference between the vaccinations was tested for a potential influence on anti-spike-RBD-IgG and spike-directed IFN-γ T cell responses at the second time point using linear models adjusting for age and sex at the first time point. Model predictions were visualized as trend lines with 95% confidence bands plotted over scatter plots between the time difference and anti-spike-RBD-IgG or spike-directed IFN-γ T cell responses.

The effects of anti-spike-RBD-IgG/spike-directed IFN-γ T cell response measurements across time as well as the effects of anti-spike-RBD-IgG on spike-directed IFN-γ T cell response at each time point were modelled in each vaccination regime using linear models. The estimated linear trends were visualized on top of scatterplots with 95% confidence bands.

The effects of age, sex, and time span between T1 and T2 on the immune response (both anti-spike-RBD-IgG and spike-directed IFN-γ T cell response) were visualized per vaccination cohort with trend lines from linear models (for age and time span) and with standard deviations (for sex).

The effect of the vaccination regime on neutralization was modelled at each time point, controlling for age, sex and time between vaccination and measurement (as well as time between T1 and T2 at the second time point) using quantile regression (due to the skewed distribution of the NI values). The effect of the vaccination regime on relative avidity was modelled at each time point, controlling for age, sex and time between vaccination and measurement (and time between T1 and T2 at the second time point) using linear models.

The significance level was set to alpha = 5% for all statistical tests. All analyses were performed with the statistic software R (version 3.6.2[1]) using the R-packages lme4 (version 1.1.21[2]) for the mixed effect regression models, lmerTest (version 3.1.0[3]) for the estimation of the Satterthwaite denominator degrees of freedom, emmeans (version 1.4[4]) for the estimation of the marginal means and the contrast tests, and quantreg (version 5.54[5]) for the quantile regression models.

1 R Core Team. R: A Language and Environment for Statistical Computing. Vienna, Austria: R Foundation for Statistical Computing, 2019 https://www.R-project.org/.

2 Bates D, Mächler M, Bolker B, Walker S. Fitting Linear Mixed-Effects Models Using lme4. J Stat Softw 2015; 67. DOI:10.18637/jss.v067.i01.

3 Kuznetsova A, Brockhoff PB, Christensen RHB. lmerTest Package: Tests in Linear Mixed Effects Models. J Stat Softw 2017; 82. DOI:10.18637/jss.v082.i13.

4 Lenth R. Emmeans: Estimated Marginal Means, Aka Least-Squares Means. 2019. https://CRAN.R-project.org/package=emmeans. (accessed Oct 14, 2021).

5 Koenker R. Quantreg: Quantile Regression. 2019. https://CRAN.Rproject.org/package=quantreg. (accessed Oct 14, 2021).
